# Validation of Kiswahili Version of WHOQOLHIVBREF questionnaire among people living with HIV/AIDS in Tanzania: a cross-sectional study

**DOI:** 10.1101/2021.01.22.21249371

**Authors:** Nuru Kondo, Tumbwene Mwansisya, Eric Aghan, Riaz Ratansi

## Abstract

**Background:** Quality of life is an important element of surveillance in people living with HIV/AIDS. WHO has developed an HIV specific quality of life tool (WHOQOLHIV-Bref) for assessing Quality of life of HIV individuals. This tool takes into account the different cultural variations that exist worldwide and hence enable assessment of the quality of life across different cultures. Despite its preliminary sound validity and reliability from several studies, the developers recommend it to be validated in different cultures to fully assess its psychometric properties before its adaptation.

**Objectives:** To evaluate the validity and reliability of WHOQOLHIV-Bref questionnaire in Tanzanian culture among people living with HIV/AIDS.

**Methods:** This was a cross-sectional study of 103 participants interviewed using a Kiswahili WHOQOLHIV-BREF questionnaire. Of, these participants 47 participants were enrolled to repeat an interview two weeks later. Internal consistency and test-retest reliability were analyzed. Validity was assessed through analysis of translational, concurrent, convergent and discriminant validity while the model performance was assessed by Exploratory and confirmatory factor analysis.

**Results:** The mean age of the participants was 40.5 ± 9.702 years. Translation validity was assessed through the WHO translational protocol and was found to be good. The internal consistency and test-retest reliability of the Kiswahili version of WHOQOL-HIV BREF were excellent: Cronbach’s alpha values of 0.89-0.90, and ICC of 0.92 p < 0.01 respectively. Concurrent valid was excellent, significant correlations were noted across all domains (correlation coefficient r > 0.3) except for physical and spiritual domains. Confirmatory factor analysis found that the six domain produced an acceptable fit to the data. The convergent and divergent validities were satisfactory.

**Conclusion:** Kiswahili WHOQOLHIV-Bref was found to be reliable and valid questionnaire among Tanzanian people living with HIV/AIDS. These findings provide support for the use of this tool in assessing the quality of life in Tanzania.

## Background

HIV/AIDS is a global epidemic with an estimated 36·9 millions of people worldwide living with HIV/AIDS(1). Sub-Saharan Africa (SSA) bears the greatest burden of HIV infections, with approximately 70% of all global estimates(1). Currently, 21.7 millions of people living with HIV worldwide are already on highly active anti-retrovirus therapy(2).

Tanzania is among the Sub-Saharan Africa countries with a high HIV burden. It has an overall HIV prevalence of 4.5%(3). It is estimated that approximately 1.5million Tanzanians are living with HIV/AIDS. Of, these estimates 1.02 million (68%) HIV individuals are already on antiretroviral therapy (ARV) coverage(3).

The introduction of highly active antiretroviral drugs in 1996 and its widespread availability, have succeeded in prolonging life by reducing mortality and morbidity related to AIDS(2). These includes the reduction of the proportion of newly infected individuals worldwide by 18% (2.1 million to 1.7million) from the year 2010 to 2018(2). The number of HIV associated deaths have also decreased from average 1.7million per year in 2010 to 770,000 in 2018 which corresponds to a 33% decline(2).

Despite this progress and benefits of the highly active antiretroviral therapy, HIV remains an incurable disease. People living with HIV/AIDS find themselves naturally facing different challenges that impair their quality of life. These challenges are attributed to HIV disease itself, stigmatization, ARV’s adverse effects and secondary comorbidities. Thus the assessment of the quality of life is an important goal in the care of HIV infected individuals(4).

QOL is a broad multidimensional concept, which addresses the general sense of wellbeing. There is no universal definition of QOL, despite various attempt to describe the concept of QOL(5). To unravel such limitation, WHO proposed a definition of QOL that could serve as a starting point to develop a thorough measure for assessing QOL. The WHOQOL Group defined QOL as “an individuals’ perception of their position in life in the context of the culture and value systems in which they live, and in relation to their goals, expectations, standards and concerns(6).”

Quality of life (QOL) in HIV disease has been widely studied for the last three decades amongst different HIV positive populations including; older patients, pediatric populations, women and military patients. The QOL in People living with HIV was also compared to the quality of life (QOL) of the HIV negative individuals(7). Various factors have been proven to strongly affect the quality of life (QOL) of HIV sero-positive individuals both in positive and negative aspect(8). These factors are Satisfaction with the health system, patient adherence to treatment, age, sex, social support, depression, longstanding illness, functional disability, symptoms severity and level of CD4+ lymphocytes counts(8).

Several tools have been used to assess the QOL of PLWHA. Majority of these tools were developed in a single culture and some of them used a poorly constructed models that omit the key aspects of QOL(9). Other tools were too long to administer and hence cumbersome in a routine busy clinic. This raised many questions regarding the generalization of the findings as well as the question of which tool will be the best to assess the QOL in a clinical environment. How such a tool would impact the current routine care of HIV patients?

To unravel this limitation, WHO developed a WHOQOLHIV-BREF to encompass a cross-cultural character and, therefore, to facilitate its use in a cross-cultural context(10). It contains 31 items that have 5 extra items that are HIV related. It is a self-administered questionnaire.

It is shorter to administer, it takes 10 minutes to complete the questionnaire. The items are grouped into 6 domains (physical. psychological, social, spiritual, level of independence and environmental domain)(11).

Several studies have been done to assess the validity and reliability of WHOQOLHIV-BREF. The results have demonstrated overall sound validity and excellent reliability(12)(13)(14). In a systematic review of HIV QOL generic and specific tools done by Vanessa et al, WHOQOLHIV-BREF was shown to be the most valid cross-cultural tool(15). It is therefore recommended as a good choice for international Assessment of quality of life in people living with HIV.

Despite the good preliminary result of WHOQOLHIV-Bref from existing studies, Vanessa et al recommended further validation studies on different cultures(15). WHOQOL team acknowledge as well further validation studies of WHOQOLHIV-Bref in new cultures, especially in SSA where the burden of HIV is high and it lacks quality data on quality of life assessment(10). Although widely used, WHOQOLHIV-Bref has not been validated in Tanzania settings.

Due to cultural variations that exist between countries WHOQOLHIV-BREF needs to be validated in different cultures to determine its psychometric properties. This questionnaire needs to be validated in Sub-Saharan settings like Tanzania where the burden is high and there is a lack of data on quality of life of HIV positive individuals. The objective of this study was to assess the validity and reliability of the Kiswahili version of WHOQOLHIV-BREF among people living with HIV/AIDS in Tanzania.

For a tool to be effective, the translation to the local language is recommended(10). Kiswahili is the national language and is used by 95% of the Tanzanians. The objective of this study was to assess the validity and reliability of the Kiswahili version of WHOQOLHIV-BREF in People living with HIV/AIDS in Tanzania.

## Methods

### Study design and setting

This was a cross-sectional design, conducted at Mnazi Mmoja, centre for treatment and care for HIV/AIDS patients.

A public health centre with approximately 12,000 HIV cohort. The centre is situated in the Dar es Salaam, a cosmopolitan city and one of the major business city of Tanzania. Dar es Salaam is one of the regions in Tanzania with a high prevalence of HIV due to its cultural diversity, tourism and economic activities.

### Study participants

Systematic random sampling was used to recruit eligible participants. All individual with a diagnosis of HIV for six months and above, age 18 and above and those who can read and write were included. The individuals with HIV and psychiatric conditions, dementia, and other cognitive diseases were excluded. Figure 1

### Questionnaire

WHOQOLHIV-Bref Kiswahili version was used for the interview after a throughout WHO translation protocol. The questionnaire was a self-administered comprised of 31 questions. Original English WHOQOLHIV-Bref questionnaire was used to assist with scoring and coding.

### Data Collection

A total of 103 PLWHA who fulfilled the inclusion and exclusion criteria were selected to take part in the study. Written informed consent was obtained from all participants. Participants were required to assess their quality of life in the recent two weeks. 48 participants were also asked to complete the Kiswahili version of WHOQOLHIV-BREF again within two weeks to measure test-retest reliability.

### Data analysis

Data entries and analyses of results were done using, the statistical Package for Social Sciences (SPSS, version 25.0) software and the analysis of moment structure (AMOS, version 26.0). Descriptive statistics of the participants were determined, categorical and numerical data were presented by frequencies and means. Internal consistency reliability was evaluated using Cronbach’s alpha for each domain and item. The intra-class correlation coefficient (ICC) and its 95% confidence interval were used for test-retest reliability.

All the variables that had negative direction questions were recoded and then screened for missing values (6%: < 20%: normal). The floor and ceiling effect were calculated and showed an overall of 2.2% and 6.0% respectively for all items (normal: <20%).

### Patient and Public Involvement (PPI)

It was not possible to involve patients or the public in the design, or conduct of this study because the idea of PPI protocol in our setting was not known. However, the dissemination of the results to the participants was pre-planned to happen through the regular HIV meetings conducted once a week within the HIV treatment Centre. All participants of this study agreed the results be made public through publication and/or presentations to bring changes without compromising the confidentiality protocol.

## Results

### Socio-demographic characteristics

Participant’s ages were ranged from 21 to 67 years, their mean age was 40.5±9.702 SD with two-third being between 45 years and below (67.0%). Majority of the participants were women (66, 64.1 %) and married individuals (42, 40.8%). 86.4% of the participants had a low level of education (did not attain tertiary education). Of the 103 participants, 71 (68.9%) were infected through heterosexual sex. A total of 85 (82.5%) participants were asymptomatic with mean time since diagnosis being 8.8years ± 6.27SD. The detailed recruitment flow diagram and socio-demographic and HIV –related characteristics of the participants are summarized in Table 1.

**Table 1:** Descriptive analysis of the baseline characteristics of the study population.

| **Variable** | **Frequency** | **Percentage** |
| --- | --- | --- |
| **Gender** |  |  |
| Male | 37 | 35.9% |
| Female | 66 | 64.1% |
| **Age (years)** |  |  |
| 18 -35 | 5 | 4.9% |
| 26 – 35 | 29 | 28.2% |
| 36 – 45 | 33 | 32.0% |
| 46 – 55 | 26 | 25.2% |
| 56 – 65 | 6 | 5.8% |
| ≥ 66 | 1 | 1.0% |
| **Marital status** |  |  |
| Single | 31 | 30.1% |
| Married | 42 | 40.8% |
| Separated/Divorced | 14 | 13.6% |
| Widow | 16 | 15.5% |
| **Education** |  |  |
| Primary | 48 | 46.6% |
| Secondary | 41 | 39.8% |
| Tertiary | 14 | 13.6% |
| **Mode of HIV- transmission** |  |  |
| Sex with male/female | 7 | 68.9% |
| Injecting drugs | 2 | 1.9% |
| Blood products | 6 | 5.8% |
| Others | 24 | 23.3% |
| **HIV symptoms status** |  |  |
| Asymptomatic | 85 | 82.5% |
| Symptomatic | 13 | 12.6% |
| AIDS | 5 | 4.9% |

### Score distribution of WHOQOLHIV-Bref

The mean scores distribution of the domains of 103 participants ranged from 10.17 ±3.18SD to 22.20 ± 4.86SD. Across domains, the environmental domain had the highest score (22.2 ±4.86). We analyzed the items with minimum and maximum scores. The overall floor and ceiling effect values were 2.2% and 6.0% respectively (values above 20% are considered significant). However the following items had a very high ceiling effect; physical pain (33.9%), HIV symptoms (33.9%), self-esteem (30.1%), non-medical dependence (28.2%), mobility/get around (27.2%), daily activities capacity (24.3%), health services availability (33.0%), stigma (40.8%), fear of the future (39.8%) and death worries (58.3%). This pattern does not lead to skewed distributions for these items. Floor effect was also detected in the item measuring sexual activity (24.3%).

### Reliability

Analysis of the 31 items showed a Cronbach alpha coefficient of 0.89 – 0.90. This result indicating that the Kiswahili version of WHOQOLHIV-Bref has acceptable internal consistency. The test-retest reliability showed a statistically significant Intra-class correlation for all items. The test-retest values were good, with the ICC ranging from 091 −0.92. (p<0.001). Table 2 shows the distribution of inter-class correlation and test-retest reliability in the facets.

**Table 2:** Distribution of Intra-class correlation and internal consistency of the Kiswahili version of WHOQOLHIV-Bref facets.

| Domains | Cronbach Alpha | Intra-class correlation | P-Value |
| --- | --- | --- | --- |
|  | (n=103) | (n=48) |  |
| Purposefulness | .892 | 0.916 | <0.001 |
| Pleasure/Enjoyment | .890 | 0.912 | <0.001 |
| Stigma | .898 | 0.919 | <0.001 |
| Fear of the Future | .896 | 0.918 | <0.001 |
| Death worries | .899 | 0.918 | <0.001 |
| Attentiveness | .895 | 0.914 | <0.001 |
| Secure feelings | .892 | 0.911 | <0.001 |
| Environmental safety | .891 | 0.911 | <0.001 |
| Energy | .889 | 0.911 | <0.001 |
| Body Appearance | .892 | 0.912 | <0.001 |
| Financial stability | .889 | 0.912 | <0.001 |
| Social involvement | .892 | 0.912 | <0.001 |
| Informative tools | .889 | 0.912 | <0.001 |
| Leisure/Recreation | .892 | 0.912 | <0.001 |
| Get around | .890 | 0.912 | <0.001 |
| Sleep | .891 | 0.911 | <0.001 |
| Daily activities | .890 | 0.910 | <0.001 |
| Work capacity | .891 | 0.911 | <0.001 |
| Self-love | .891 | 0.912 | <0.001 |
| Personal relationships | .892 | 0.912 | <0.001 |
| Sex life | .896 | 0.916 | <0.001 |
| Friends/Relative Support | .888 | 0.911 | <0.001 |
| Living Condition | .889 | 0.910 | <0.001 |
| Health Services | .892 | 0.911 | <0.001 |
| Transport System | .890 | 0.914 | <0.001 |
| Negative feelings | .895 | 0.911 | <0.001 |

### Validity

#### Construct validity

The six domain WHOQOLHIVBREF model was assessed by confirmatory factor analysis (CFA) using the AMOS software to examine whether it explains the relationships among domains and facets. Majority of the WHOQOLHIVBREF items produced substantial factor loadings and analysis of model fit produced an acceptable fit to the model (X2= 658.319, df =362, RMSEA= 0.09).

#### Concurrent validity

WHOQOLHIVBREF had a moderate correlation with three self-evaluated general questions/items (the overall quality of life, general health perception, and self-perceived health status). It was found that scores of four domains (psychological, social, environment and level of independence) were positively correlated with the three self-evaluated general questions. Their range of Pearson’s correlation coefficient within the domains were statistically significant and were above 0.3 which is recommended for evaluating concurrent validity. Table 3 shows the correlation between each domain scores with the three self-evaluated general questions.

**Table 3:** Correlation between Kiswahili version of WHOQOLHIV-Bref and general health measures.

| **Domain** | **Self-evaluated QOL** | **Self-evaluated**<br>**General Health** | **General health** |
| --- | --- | --- | --- |
| **Physical** | 0.13 | -0.04 | -0.07 |
| **Psychological** | 0.373*** | 0.22** | 0.52*** |
| **Environmental** | 0.29** | 0.20** | 0.32** |
| **Social** | 0.37*** | 0.38** | 0.34** |
| **Level of independence** | 0.44*** | 0.21** | 0.34** |
| **Spiritual** | 0.03 | -0.60 | -0.15 |

\*\*p-value 0.01-0.05 \*\*\* p-value < 0.001

#### Convergent Validity

Convergent validity was determined by the correlation between items and their respective domains. All items showed moderate to strong correlations with their respective domain and r coefficients ranged from 0.243 to 0.762 (p<0.01). The highest correlations of items were seen in the social domain where r coefficients ranged from 0.62 −0.76. The overall convergent validity was good. Table 4 shows the correlation between items and their respective domains.

**Table 4:** The Pearson r correlation between items and their respective domains.

|  | ITEM-DOMAIN |  | ITEM-DOMAIN |
| --- | --- | --- | --- |
|  | CORRELATION |  | CORRELATION |
| **Physical domain** |  | **Spiritual Domain** |  |
| Physical pain | -0.485** | Stigma | -0.741** |
| HIV Symptoms | -0.596** | Fear of the Future | -0.627** |
| Energy | -0.477** | Death worries | -0.579** |
| Sleep | 0.48** | Enjoyment | -0.058** |
| **Social domain** |  | **Independent Domain** |  |
| Sexual activities | 0.616** | Medical dependence | -0.340** |
| Personal relationship | 0.708** | Mobility | 0.676** |
| Friend support | 0.762** | Daily activities | 0.641** |
| Social inclusion | 0.656** | Work Capacity | 0.665** |
| **Environmental Domain** |  | **Psychological Domain** |  |
| Secure feelings | 0.615** | Attentiveness | 0.608** |
| Environmental safety | 0.638** | Purposefulness | 0.409** |
| Financial stability | 0.665** | Body appearance | 0.568** |
| Informative tools | 0.703** | Negative feelings | -0.243** |
| Leisure | 0.669** | Self-Love | 0.667** |
| Living condition | 0.538** |  |  |
| Health services | 0.563** |  |  |
| Transport system | 0.669** |  |  |

#### Discriminant validity

Concerning discriminant validity, spiritual and physical domain were highly discriminated from the rest of the domains. Their correlation coefficients were low compared to their squared root of average variance extracted (AVE). The other four domains were poorly discriminated, with their correlation coefficients greater than their squared root of AVE. This results concluded that the discriminant validity was satisfactory when compared to the three domains founds in the exploratory factor analysis of Kiswahili WHOQOLHIV-BREF.

## Discussion

The results of this study suggested that the Kiswahili version of WHOQOL-HIV BREF is a valid and reliable instrument for evaluation of the quality of life in PLWHA. In general, the internal consistency and test-retest reliability of the Kiswahili version of WHOQOL-HIV BREF was excellent. The construct validity of the questionnaire measured by major types of validity evidence (translational, concurrent, convergent, and discriminant) provides strong valid results supporting its use in quality of life screening of PLWHA.

The mean age of our study participants was 40.5 ± 9.7SD, which is similar to the Georgia and Portugal validation studies(16)(17). The vast majority of the participants were female and heterosexual route was the predominant mode of HIV transmission this is similar to Ethiopian study(13). This could be related to religious and cultural influence in the country that favours the heterosexual route. The predominance of the female is explained by the inability to negotiate safe sex among African women and the cultural aspects of early marriages. In other studies, the majority of the participants were male and homosexual route was the predominant mode of HIV transmission(12)(17)(18).

Descriptive analysis of the score showed a ceiling effect on some of the items. It was noted that the scores on these items indicated the most favourable circumstance for the respondents and it can be attributed to the religious belief around the items in question. This ceiling pattern did not affect the normality of the distribution for these items that warrant the use of the non-parametrical method. Floor effect was detected in the item measuring a sexual activity and this is due to the cultural difficulty in revealing one’s sexual practice. Other studies also encountered floor and ceiling effects in different items according to their cultural context(12)(14)(16).

Regarding the reliability of the Kiswahili version of WHOQOL-HIV BREF. This present study demonstrated an excellent internal consistency of the tool. Test-retest reliability of the WHOQOLHIVBREF also was excellent hence make the questionnaire very reliable. These findings were similar to the majority of the studies that were conducted in other cultures as well(13)(14)(16)(17)(18).

The Kiswahili WHOQOLHIVBREF questionnaire was found to be valid. This was deducted through analysis of Translational validity, concurrent validity, convergent and discriminant validity which showed a good construct validity. These results were similar to other studies previously done using language translated versions of WHOQOLHIVBREF questionnaire(14)(16)(17)(18).

Quality of life measured by an individual through three self-evaluated general questions showed a high correlation with the scores in the domains except for physical and spiritual domain. This reflects the convergence of the construct towards the quality of life outcome. The low correlation of the physical and spiritual domain has been also reported in Malaysia and Taiwan studies(14)(18). The two possible explanation for these findings are; first, the presence of overlapping constructs between physical and spiritual which failed to discriminate the items leading to different interpretation/perceptions. Second, studies have indicated that religion and culture can have an influence on the lifestyle and shapes the experiences of illness, pain, and end-of-life care. Majority of Tanzanians are likely to have religious beliefs that are associated with poor medical seeking behaviours(14)(18)(19).

Convergent validity of Kiswahili WHOQOLHIVBREF was found to be excellent. The social domain had the highest correlation among the six domains this highlight the role of a good social support system in enhancing the quality of life of an individual. The discriminant validity was satisfactory. These findings were observed in other studies as well(19).

### Study limitation

The sensitivity to change of the Kiswahili WHOQOLHIVBREF was not assessed due to the cross-sectional design used. Longitudinal studies can help to answer the responsiveness of the questionnaire to clinical stages of HIV/AIDS.

## Conclusion

The WHOQOL-HIV BREF questionnaire revealed excellent reliability and validity among Tanzanian’s people living with HIV/AIDS. These findings provide evidence to support the use of the WHOQOL-HIV BREF as a tool of QOL screening in HIV positive individuals in Tanzania. Data on quality of life can be obtained using this questionnaire and can help us to target different needs that are arising in HIV populations. This can help in resources allocation, to device new interventions targeting treatment and amelioration of quality of life and others.

## Supporting information

https://www.equator-network.org/reporting-guidelines/strobe/

## Data Availability

Data are protected and not available to the public for confidential purposes.

## Authors contributions

All the authors participated in the study protocol, design and review of the final manuscript.

## Ethical Approval

This study was reviewed and approved by the Family Medicine Dissertation Committee, The Aga Khan University Research committee (AKU-RC) and The Aga Khan University Ethics Review Committee (AKU-ERC) before study commencement. Ethical consideration: Permission for data collection from the Mnazi Mmoja hospital was given by District Medical Officer (DMO) research board of Ilala Municipal.

## Transparency declaration

I, Nuru Kondo, the lead author, affirms that this manuscript is an honest, accurate, and transparent account of the study being reported; that no important aspects of the study have been omitted; and that any discrepancies from the study as planned (and, if relevant, registered) have been explained.

## Data dissemination

The results of this study was disseminated to the board of graduate studies of Aga Khan University in partial fulfilment of the requirements for the degree of Master of Medicine in Family Medicine. The dissemination of the results to the participants was hindered due to COVID19 pandemic and social distancing policy.

## Acknowledgment

Authors wish to thanks all the participants of this study.

